# Mobile phone communication for improving uptake of antiretroviral therapy in HIV-infected pregnant women: updated systematic review and meta-analysis

**DOI:** 10.1101/2021.12.18.21267982

**Authors:** Jacques L. Tamuzi, Gomer Lulendo, Patrick Mbuesse, Thierry Ntambwe

## Abstract

**Objective:** The aim of this systematic review is to provide some evidence on the use of mobile phone communication for improving ARV adherence during pregnancy, as well as to investigate whether text messaging on mobile phones could improve follow-up in HIV-exposed infants.

**Methods:** We did a systematic review and meta-analysis, using CENTRAL (Cochrane Central Register of Controlled Trials), Scopus, MEDLINE via PubMed, Web of Science, and CINAHL to search for studies in English published between 5 may 2016 to May 2021 that assessed the effects of mobile phone in HIV-infected pregnant women. We used MetaPro version 3.0 to compute the OR and RR and their 95%CI. We performed random-effects model meta-analysis for estimating pooled outcomes.

**Results:** Nine studies were included in the meta-analysis. The pooled maternal post-partum retention was (OR 2.20, 95%CI: 1.55 – 3.13, I^2^ = 53.20%, P < 0.001).

In the same line, the pooled odds of ART uptake was (OR 1.5, 95%CI: 1.07-2.11, I^2^ =0%, P = 0.020) and we found statistically significant impact of mobile phone on HIV testing at 6 weeks and above among HIV exposed children (OR 1.89, 95%CI: 1.04 – 3. 48, I^2^ = OR 1.89, 95%CI: 1.04 – 3. 48, I^2^ =88.04%, P = 0.032).

**Conclusion:** In comparison to our previous review, this updated review focuses on moderate evidence for mobile phone communication in HIV-infected pregnant women. The results showed that using a mobile phone improved maternal post-partum retention, ART uptake, and infant HIV testing at 6 weeks and older.

## 1. Background

In 2020, there were 37.7 million HIV-positive people worldwide, with women and girls accounting for 53% of all HIV-positive people, and children accounting for 1.7 million [1]. While progress has been made in these priority countries, much more effort is required to meet the Global Plan’s target of 90 percent reduction in new infections among children [2]. Every week, approximately 5000 young women between the ages of 15 and 24 become infected with HIV [1]. Six out of every seven new HIV infections among adolescents aged 15–19 years in Sub-Saharan Africa are among girls [3], young women aged 15–24 years are twice as likely as men to be HIV positive. In 2020, approximately 4200 adolescent girls and young women aged 15–24 years were infected with HIV every week [3]. The most important factor in preventing perinatal transmission in HIV-positive women is antiretroviral therapy (ART) during pregnancy [4]. Without treatment, the risk of perinatal transmission is 25%, which drops to 1.8 percent with zidovudine alone and 0.5% with ART treatment regimens [4]. Individuals living with HIV should begin ART and achieve viral suppression prior to pregnancy, regardless of method of conception [5]. Full viral suppression for at least three months prior to conception is recommended, or at least two viral load measurements below the level of detection at least one month apart [6]. Several large studies have found that suppressing the plasma HIV viral load reduces the risk of sexual transmission [6–8]. Indeed, new maternal HIV infection among pregnant women significantly contributes to mother–to–child HIV transmission (MTCT) [9–11]. According to UNAIDS 2020, new maternal HIV infection during pregnancy was the second leading cause of perinatal HIV transmission in 2019 [3].

Pregnancy is a high-risk period for HIV acquisition [12]. Thomson et al. found that the per-act probability of HIV acquisition increased by 2.8 during pregnancy compared to non-pregnancy [12]. The pooled HIV incidence among pregnant women in Sub–Saharan African and other countries was reported to be 2.1 per 100–person years in a meta–analysis of studies conducted between 2014 and 2018 [13]. The majority of HIV-infected children are infected through MTCT, which can occur during pregnancy, labour and delivery, or breastfeeding. In the absence of any intervention, the risk of such transmission in non-breastfeeding populations is 15–30% [14]. Breastfeeding by an infected mother raises the risk by 5–20%, for a total of 20–45% [14]. Preterm birth, prolonged rupture of membranes, concurrent syphilis, chorioamnionitis, maternal viral load >100 000 copies/mL, and obstetrical interventions, including elective caesarean delivery, are all risk factors for perinatal transmission [14, 15]. However, among women with undetectable viral loads, the length of time between membrane rupture and birth may not be associated with an increased risk of perinatal transmission [16]. When compared to women with chronic and well-managed HIV, acquiring HIV during pregnancy or lactation increases the risk of perinatal transmission by 15 and 4-fold, respectively [17–20]. In newly acquired maternal HIV infection, starting ART as soon as possible after infection can reduce maternal HIV viral load and the risk of MTCT [21]. ART reduces the risk of HIV transmission from mother to child [22–24]. Increased access to ART during pregnancy and breastfeeding has had the greatest impact on reducing perinatal HIV-1 transmission. According to UNAIDS, active management of pregnant women living with HIV-1 has reduced new HIV-1 infections among children by half, from 310 000 in 2010 to 150 000 in 2019 [1]. Despite the evidence showing pregnant women having the highest level of adherence to ART [25], a low level of adherence has been reported in some settings among pregnant women [26]. In fact, the adherence rate among pregnant women varies across different settings (within and across countries) from 35% to 93.5% [27]. A study reported low adherence levels among pregnant women in rural Kwazulu Natal, South Africa [26]; however, a high level of adherence (90%) was reported in Kenya [28]. An adherence level of 73% during pregnancy was reported in Malawi, but dropped to 66% by three month postpartum [26].

Reviewing the literature, the field of HIV adherence interventions is quite broad. Major reviews of adherence interventions, primarily conducted in developed countries, revealed that the most effective are typically patient-centered, behavioral interventions designed to increase patient knowledge and efficacy through practical medication management skills [29–33]. While some social and behavioral scientists have become heavily involved in developing adherence measures, studying explanatory factors, and developing programs to improve patient adherence, others [33–36] have argued, l’arbre qui cache la forêt’ [33, 36], meaning patient adherence is multidisciplinary and complex. Coomes et al. proposed a conceptual model that integrates short message services (SMS) communication functionality with important psychosocial factors that could mediate the impact of SMS communication on health outcomes [33, 37]. This model is based on limited but growing research that has used SMS to assist patients in self-management or adherence across a variety of disease conditions [33, 37]. The use of mobile phones to improve HIV-related health outcomes is being emphasized in most HIV-affected countries, as emerging evidence suggests that reminder SMS can increase adherence to ART and retention in care, decrease viral load and treatment interruptions, and improve communication with healthcare personnel [38]. The main aim of this systematic review is to provide some evidence on the use of mobile phone communication for improving ART adherence during pregnancy, as well as to investigate whether mobile phone text messaging could improve follow-up in HIV-exposed infants.

## 2. Methods

This systematic review and meta-analysis was updated following the Preferred Reporting Items for Systematic Reviews and Meta-analyses (PRISMA) guidelines [39]. The study protocol was registered in Prospero: CRD42016032800. The previous review version was published [33].

### 2.1. Data sources and searches

From 5 may 2016 to May 2021, we searched the following electronic databases: CENTRAL (Cochrane Central Register of Controlled Trials), Scopus, MEDLINE via PubMed, Web of Science and CINAHL (EBSCO). Both text words and Medical subject heading (MeSH) terms, for example pregnant women, adherence, antiretroviral, antiretroviral therapy, HAART, HIV, acquired immunodeficiency syndrome, SMS, texting, text message, short message service, cell phone, phone, telephone, mobile health, mHealth, mobile phone, short message and randomized controlled trial were used in the search strings. Furthermore, we used Boolean search in different combinations with the adaptation of the literature search strategy that will be convenient to each database. We suggested the following search strategy: (Pregnant women) OR (pregnant) OR (pregnancy) OR (pregnancies) OR (gestation) OR (mother to child transmission) OR (vertical transmission) OR (mtct) OR (pmtct) OR (perinatal transmission) AND (Cellular phone) OR (telephone) OR (mobile phone) OR (text message*) OR (testing) OR (short message*) OR (cell phones) OR (SMS) OR (short message service) OR (text) OR (mobile health) OR (telemedicine) OR (health) OR (health communication) OR (health education) OR (behavior) OR (ehealth) AND (Antiretroviral therapy) OR (anti-retroviral agents) OR (antiretroviral) OR (ART) OR HAART AND (Randomized controlled trial) OR (controlled clinical trial) OR (randomized controlled trials) OR (random allocation) OR (double-blind method) OR (single blind method) OR (clinical trial) OR (trial) OR (clinical trials) OR (clinical trial) OR (single* OR double*) OR (treble* OR triple*) AND (mask* OR blind*) OR (placebos) OR (placebo*) OR (random*).

We also conducted an electronic search for potentially eligible studies through conferences websites: International AIDS conferences, The European AIDS Clinical Society (EACS) conferences, International AIDS Society Conference on HIV Pathogenesis and Treatment (IAS). We also searched for unpublished and ongoing studies in prospective clinical trial registries such as ClinicalTrials.gov and WHO International Clinical Trials Registry Platform.

### 2.2. Eligibility criteria

To include eligible studies, we used the PICOS (P: population, I: intervention, C: comparators, O: outcome, S: study design) approach. I P: HIV-infected pregnant women who have been assigned to the intervention. (ii) I: communication via mobile phone (iii) C: HIV-infected pregnant women who do not receive treatment (iv) O: ANC visits, post-partum visits, ART enrolment, and HIV testing for children. The polymerase chain reaction (PCR) was used to test children for HIV. S: intervention studies that looked at the impact of mobile phone communication on HIV-infected pregnant women. We excluded studies for the following reasons: I qualitative studies that reported mobile phone use in HIV-infected pregnant women, (ii) intervention studies that did not report review outcomes, and (iii) non-primary studies such as reviews. We looked at randomized controlled trials (RCTs) that looked at the efficacy of mobile phone communication for improving therapy in HIV-infected pregnant women who were taking ARVs or were about to start Highly active antiretroviral therapy (HAART). The interventions included daily, weekly, or monthly SMS reminders or other forms of mobile phone communication aimed at reminding the patient about ARV adherence, with the patient expected to respond within 48 hours, the clinician to respond, non-responders called and calls, and the comparisons were standard care or no mobile phone communication. Maternal outcomes were the initiation of antiretroviral therapy, maternal clinical attendance (ante and postpartum), and infant outcomes were mother to child transmission of HIV and number of infants tested for HIV at 6 weeks.

### 2.3. Study selection and data extraction

All studies that were potentially relevant for inclusion and were retrieved using databases and manual hand searching were stored in EndNote X 8 for management. After automatically excluding duplicate studies, the titles and abstracts of the remaining studies were screened. Second, we removed titles and abstracts that appeared to be irrelevant to inclusion. Third, after deduplication and title and abstract screening, potentially relevant studies were reviewed for full-text (where available), and ineligible studies were removed. Fourth, the literature in the full-texts was filtered using pre-defined criteria.

After the selection of eligible studies, the following data were extracted using a pre-prepared excel sheet: trial characteristics: trial design, the risk of bias assessment, the number of participants included and excluded length of follow-up (in weeks) and lost to follow-up (number of patients); participant characteristics: country of origin, sample size, setting, age, date of the study, the range of gestational age, ante or postpartum period, comparison group; intervention characteristics: duration of intervention (in weeks, intensity per week and total time expressed in hours); type of intervention: e.g. SMS or mobile phone calls; outcomes: primary and secondary outcomes relevant to this review. Two authors (JLT and PB) independently conducted study selection and data extraction. Any inconsistency or disagreement between the two authors was resolved by consensus.

The methodology used for collecting and analysing data was based on the guidance of the Cochrane Handbook of Systematic Reviews of Interventions [40]. Authors (JT and PB), working independently, reviewed the abstracts of all studies identified through database searches or other resources. Where there was any question of eligibility, we obtained the full text of the article for closer examination. We used a data extraction sheet to capture data and entered the data into ProMeta software version 3.0.

### 2.4. Quality assessment

Bias risk assessment in included studies using the Cochrane risk of bias tool, we independently assessed the risk of bias [40]. The recommended approach for assessing trial quality in Cochrane Reviews studies is based on six domains (sequence generation, allocation concealment, blinding of participants, personnel and outcomes, incomplete outcome data, selective reporting and other issues). The tool’s first step required describing what was reported to have occurred in the study. The tool’s second step involved assigning a risk of bias judgment for that entry based on whether it was high, low, or unclear. If a disagreement could not be resolved, the third author was consulted to make a final decision.

### 2.5. Data analysis

ProMeta version 3.0 was used for all statistical analyses. With 95% CI, we used odds (ORs) ratio and risk ratio (RR) as the primary categorical outcome measures of effect. The results were displayed in the form of forest plots. We used ORs and RR and their 95%CI to examine the degree of association between antenatal and postnatal visits, ART uptake among HIV-infected pregnant women, and HIV testing at 6 weeks among HIV-exposed children. The random-effects model was used to calculate the overall pooled ORs and RR estimates. The inconsistency index (I^2^ statistics) was used to assess the magnitude of heterogeneity among included studies, with I^2^ values greater than 25%, 25–75%, and 75% indicating low, moderate, and high heterogeneity, respectively [40]. We used Egger’s regression test [41] and Begg and Mazumdar’s rank correlation test [42] to assess publication bias. The effect of interventions and standard care groups was weighted using subgroup analysis in both individually randomized trials and cluster randomized trials. Because we included binary outcomes of CRTs and IRTs, we can safely pool them in meta-analysis due to the lack of systematic differences in effect estimates [43]. Subgroup analysis was performed in case of high heterogeneity. Furthermore, we performed sensitivity analysis to ensure that the overall pooled estimate was not influenced by a single study.

## 3. Results

Figure 1 depicts electronic data sources and the selection process. Initially, CENTRAL (Cochrane Central Register of Controlled Trials), Scopus, MEDLINE via PubMed, Web of Science, and CINAHL (EBSCO) were used to identify a total of 2020 records. After removing 98 duplicate records, the remaining 1922 studies were screened based on titles and abstracts, and the remaining 1894 records were excluded. 72 of the 79 studies’ titles and abstracts were unrelated to the study’s objectives. The remaining seven studies were screened for inclusion in the analysis by critically reading their full-texts. Also, additional two eligible studies were included from the previous systematic review. In the end, a total of 9 studies were included in this meta-analysis.

**Figure 1:**
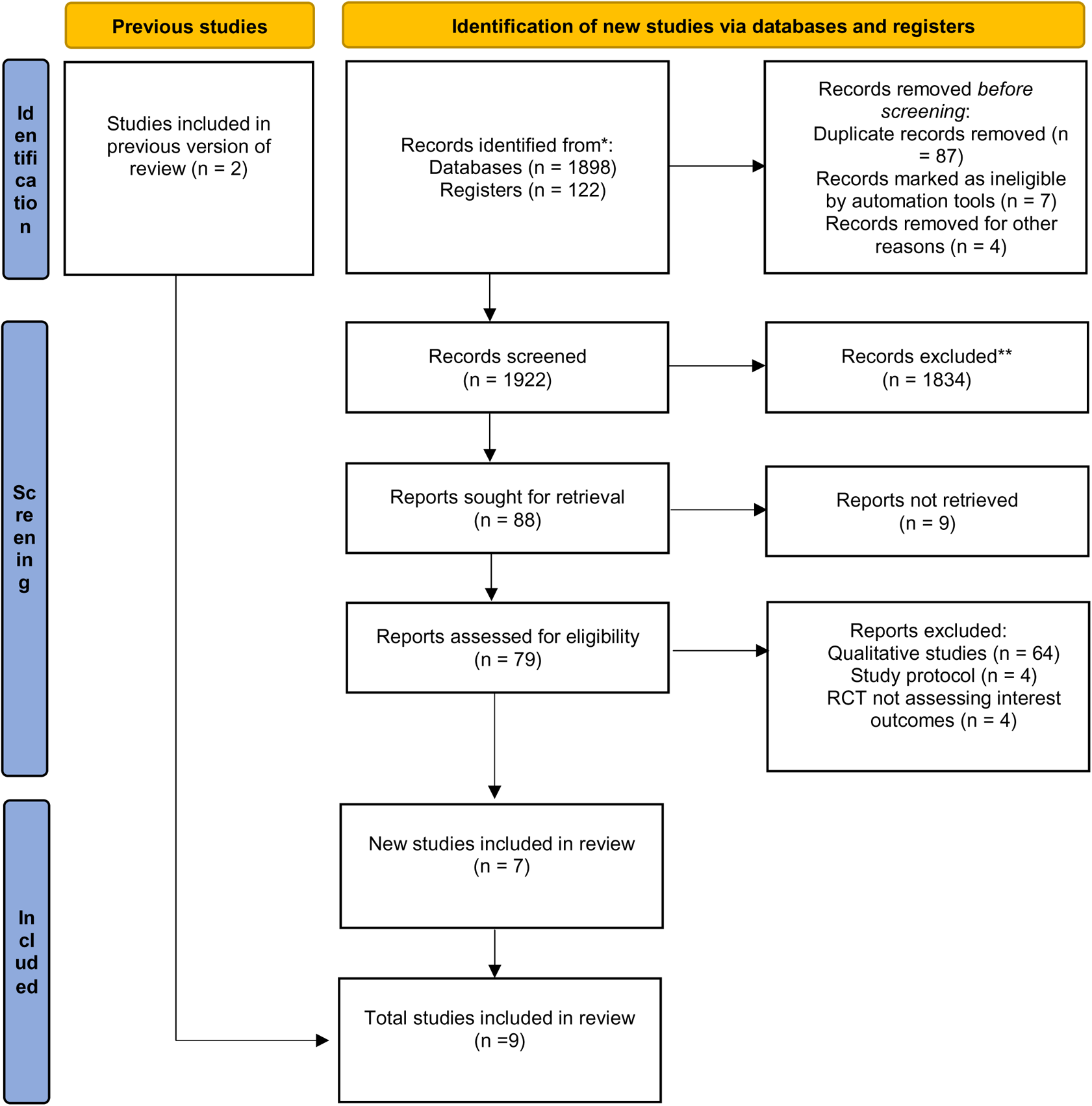
Flow diagram of included studies. For more information, visit: http://www.prisma-statement.org/

### 3.1 Characteristics of included studies

Table 1 shows the characteristics of the nine eligible studies included in this systematic review and meta-analysis. All of the included studies were carried out in five Sub-Saharan African countries: Kenya (n = 5) [44–48], Malawi (n = 1) [49], South Africa (n = 1) [50], Botswana (n = 1) [51], and Uganda (n = 1) [52]. The sample size of studies that are eligible ranges from 165 to 2472. All of the included studies took place between 2014 and 2021. The age range ranged from 16 to 29.2 years. The gestational age range is approximately 14 to 36 weeks. The interventions differed between studies. Two studies found that women sent up to 14 text messages during pregnancy and after giving birth. The message content and schedule were the same as in the previous study that demonstrated the efficacy of this intervention [46, 47]. Another study sent an automated SMS every four weeks from the second to the twelfth week [51], and one study assigned 3 to 6 SMS messages each week at a self-selected time of day and in their preferred language [44]. Kinuthia et al. assigned interventions such as 1-way SMS (participants received SMS but were unable to send messages to the study) and 2-way SMS (participants received SMS but were unable to send messages to the study) [45], to studies that did not specify the number of SMS reminders [49, 52]. Another intervention consists of two phone calls during the first week of starting PMTCT services, followed by one call per week until the participant delivers (a maximum of 26 calls), two calls during the first week after delivery, and one call per week for the next 14 weeks (a maximum of 16 calls) [48]. The risk of bias assessment of included 9 studies as showed in the Figure 2. Five studies reported a low risk of random sequence generation [44-46, 48, 51], two studies reported an unclear risk [49, 52], and two study reported a high risk [47, 50]. Two studies had a low risk of allocation concealment [45, 46], six studies had an unclear allocation concealment [44, 47–50, 52], and one study had a high risk [51]. Two studies [45, 51] had low risk of participant and personnel blinding, four studies were unblinded [46, 48, 50, 52], and three studies were unclear [44, 47, 49]. All of the included studies reported a low risk of outcome assessment blinding. Only one study reported a high risk of an incomplete outcome, while the other studies reported a low risk. Reporting bias was minimized in all studies, and four studies reported low risk of other bias [44–46, 48], while other studies reported high risk of other bias (recall bias, different in some baseline characteristics, short intervention duration, blunted the effect of the interventions on retention and null effect of the trial) [47, 49–52].

**Figure 2:**
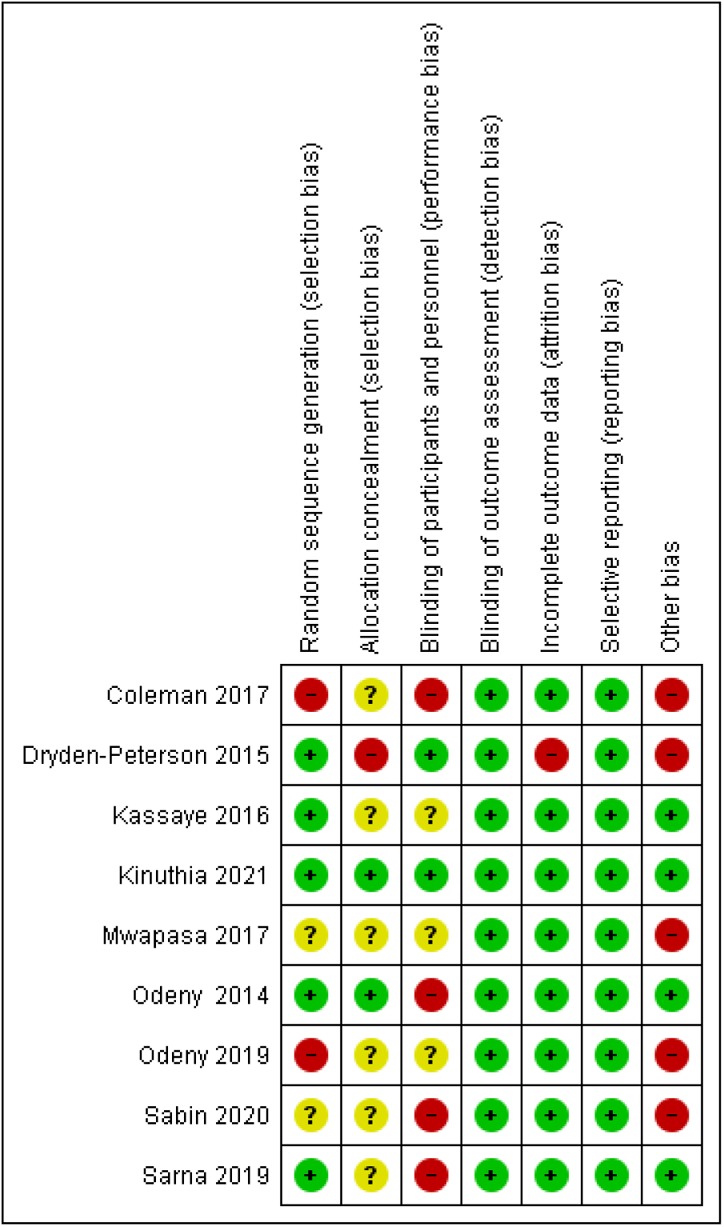
risk of bias summary

**Table 1:** Characteristics of included studies in the review

| Study ID/Setting | Methods | Participants | Interventions | Outcomes |
| --- | --- | --- | --- | --- |
| **Coleman 2017**<br>**South Africa** | non-blinded, non-randomized retrospective intervention | All participants were HIV-positive women who attended their first ANC visit between 1 April 2013. 235 HIV-infected pregnant women twice per week in pregnancy and continued until the infant's first birthday. | Maternal health SMSs were sent to HIV-infected pregnant women twice per week in pregnancy and continued until the infant's first birthday, compared to a control group of HIV-infected pregnant women who received no SMS intervention. | **At least 4 ANC visits**<br>Intervention= 60 /73<br>Control= 55/ 94<br>**PCR After six weeks**<br>Intervention= 36/ 192<br>Control= 110/447 |
| **Dryden-Peterson 2015**<br>**Botswana** | stepped-wedge cluster randomized trial | 366 HIV-infected pregnant women enrolled in PMTCT. 169 in intervention group and 156 in control group (analyzed for CD4 before 26 weeks' gestation). 154 in intervention group and 153 in control group (analyzed for ART before 30 weeks' gestation). The median gestational age at antenatal clinic registration was 18 weeks. Median(range): 28 (25-33) years. | An automated sending SMS every 4 weeks from the second week until the twelfth week. A 1-2-hour participatory session for clinic staff. Longitudinal support by study team member to antenatal clinics. | **ART initiation before 30 weeks' gestation**<br>Intervention group: 56/154<br>Control group: 37/153<br>**Impact on specific Elements of the prevention of MTCT cascade**<br>Intervention group: 168/169<br>Control group: 155/156 |
| **Kassaye 2016**<br>**Kenya** | Cluster-Randomized Controlled Study | A total of 550 HIV-infected pregnant women were enrolled into the study from June 2012 to June 2013, 280 in the intervention arm and 270 in the control arm. Women were eligible to enrol in the study if they were less than 32 weeks of gestational age, were not currently receiving antiretroviral therapy. Median age (IQR) 25.6 (22, 29) years. | A mobile web-based communications software system allowed for semi-automated delivery of preloaded messages. Participants received 3 to 6 SMS messages each week at a self-selected time of day and in their preferred language. | **Maternal Uptake of Antiretroviral Medications**<br>Antiretrovirals were given to 243/280<br>Control: 226/270<br>**Facility-Based Delivery and Infant Uptake of Antiretrovirals.**<br>Intervention: 142/280<br>Control: 134/270 |
|  |  |  |  | **Infant antiretroviral receipt increased by 6 weeks postpartum**<br>Intervention: 279/242<br>control arm: 264/228.<br>**Infant HIV testing at 6 weeks of age**<br>intervention arm: 242/243<br>control arm: 228/240<br>**HIV transmission (positive HIV DNA PCR test) at 6 weeks**<br>Intervention: 1 /242<br>three infants 4 /228 |
| **Kinuthia 2021 Kenya** | randomized nonblinded 3-armed trial | 900 eligible HIV infected pregnant women, 824 (91.6%) were enrolled and randomized, 271 to 1-way SMS, 276 to 2-way SMS, and 277 to the control arm. Median age was 27 years (interquartile range (IQR) 23 to 31), and median gestational age was 24.3 weeks (IQR 18.3 to 29.6). | Participants were individually randomized by study nurses 1:1:1 to 1 of 3 arms: control (no SMS), 1-way SMS (participants received SMS but were unable to send messages to the study), and 2-way SMS (participants received SMS and could message the study). | **Maternal virologic nonsuppression**<br>Two ways:22/249<br>One way:26.32/235<br>Control: 24/243<br>**On-time visit attendance**<br>Two ways: 245/276<br>One way:241/271<br>Control: 226/256<br>**Infant HIV infection or death.**<br>Two ways:20/252<br>One way:11/240<br>Control: 13/252 |
|  |  |  |  | **Maternal ART adherence**<br>**≥95%**<br>Two ways: 197/276<br>One way: 191/271<br>Control: 181/256 |
| **Mwapasa 2017**<br>**Malawi** | Cluster randomised control trial | 1350 HIV infected pregnant women. Median age (IQR): 29.2 (24.8 – 33.3) | Mother–infant pair clinics, which deliver integrated HIV and non-HIV services, mother–infant pair clinics plus electronic text message (SMS) reminders for mother–infant pairs who miss scheduled eMTCT follow-up clinics, and current standard of care. | Retention of HEIs at 12-Month Postpartum<br>MIP + SMS: 78/399<br>SOC: 24/ 300<br>Maternal Retention at 12 Months<br>MIP + SMS: 268/399<br>SOC: 208/300<br>infant retention at 12 months<br>MIP + SMS: 323/399<br>SOC: 234/300 |
| **Odeny 2014**<br>**Kenya** | Parallel-group, unblinded, randomised controlled trial. | 388 HIV-infected pregnant women enrolled in PMTCT. 195 women were in SMS and 193 in control group after randomisation. HIV-positive pregnant women at least 18 years old, Gestation >28 weeks. | The intervention consisted of 14 messages. Up to 8 were sent during pregnancy (weeks 28, 30, 32, 34, 36, 38, 39, and 40). Additional messages were sent weekly for the first 6 weeks after delivery. | **Maternal clinic attendance (8 weeks' post-partum)**<br>SMS group: 38/194<br>Control group: 22/187<br>**Infant HIV DBS testing (8 weeks' post-partum)**<br>SMS group: 172/187<br>Control group: 154/181 |
| **Odeny 2019**<br>**Kenya** | cluster randomized, stepped-wedge trial with 2 observation time periods | Clusters were defined as public health facilities supported by the Family AIDS Care and Educational Services (FACES) program to provide PMTCT services in Kenya. The median age was 27 years (interquartile range [IQR] 23–30). The median gestational age was 30 | Women registered in the SMS system received up to 14 text messages during pregnancy and after delivery. The message content and schedule were the same as those used in the earlier study demonstrating the efficacy of this intervention. | **Infant HIV testing**<br>SMS group: 1,466/1,613<br>(Control group: 609/713)<br>**Maternal postpartum retention**<br>SMS group: 1,548/1,725<br>(Control group: 571/747) |
|  |  | weeks (IQR 28–34). |  |  |
| **Sarna 2019**<br>**Kenya** | Parallel-group, unblinded, randomised controlled study | 404 pregnant women living with HIV who were between 14 and 36 weeks of gestation, aged ≥16 years, residing in Kisumu(Kenya) and planning to stay there for the next 12 months. | The sessions were structured to consist of 2 phone calls during the first week of starting PMTCT services, followed by 1 call/week until the participant delivered (maximum of 26 calls), followed by 2 calls during the first week after delivery and 1 call/week for 14 weeks thereafter (maximum of 16 calls). | **Received complete ANC package**<br>Intervention:192/207<br>Control:181/196<br>**Attended 3 PNC visits per national protocol**<br>Intervention:42/207<br>Control:25/197<br>**Attended 6-week PNC visit**<br>Intervention:170/207<br>Control:139/197<br>**PCR testing at 6 weeks**<br>Intervention: 168/ 181<br>Control: 87/ 127 |
| **Sabin 2020**<br>**Uganda** | Randomized Controlled Trial | A total of 165 HIV-positive women initiating ART were enrolled between June 2015 and January 2016. The average age was 25 years, with mean gestational age of approximately 21 weeks. | Intervention participants selected a text message from 10 options in consultation with a study coordinator. The message options had been developed during an earlier phase of the study with input from HIV-positive mothers and healthcare providers who conduct counseling with PPPW. | **Visit Retention**<br>Intervention: 56/67<br>Control: 56/64<br>**Postpartum Retention at Month 3**<br>Intervention: 54/67<br>Control: 52/64 |

### 3.2. Publication bias assessment

We assessed quantitatively publication bias with Egger’s regression test and Begg’s correlation test. Both Egger’s regression test (t = 0.97, P = 0.404) and Begg and Mazumdar’s rank correlation test (z = 0.49, P = 0.624) for ANC visits outcome. HIV testing at 6 weeks and above revealed the Egger test of t = 0.92 and P = 0.424; and Begg and Mazumdar’s rank correlation test (z = 1.47, P = 0.142). This results showed the absence of publication bias.

### 3.3. Pooled odds and relative ratios estimate

We included nine studies to explore the impact of mobile phone communication in improving the couple HIV-infected pregnant women and children outcomes. The pooled RR estimate (RR = 1.02, 95%CI: 0.95 – 1.10, I^2^ = 65.65%, P = 0.565) showed that the odds of antenatal visits in HIV-infected pregnant women receiving mobile phone interventions was not statistically significant compared to non-mobile phone group (Fig 3).

**Figure 3:**
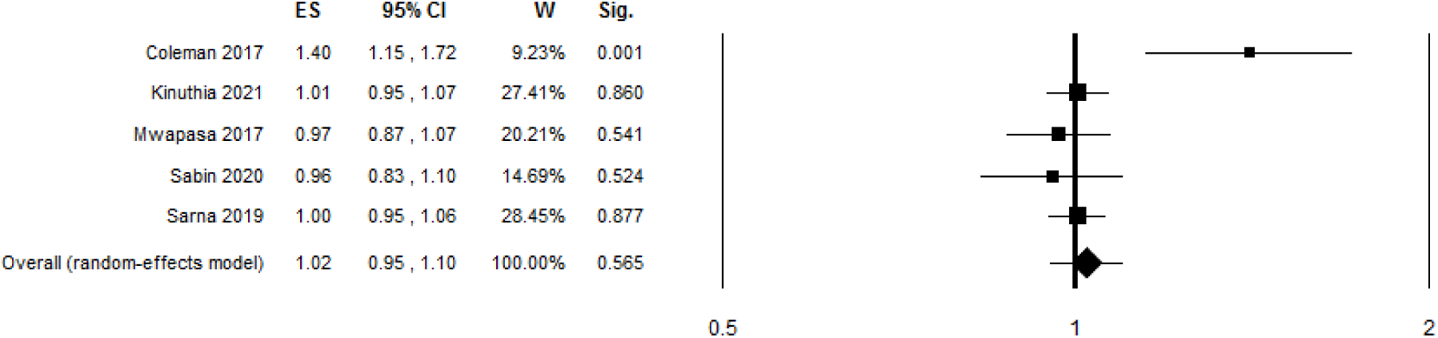
Forest plot of ANC visits among HIV-infected pregnant women: mobile phone vs control groups

Pooled odds of Maternal post-partum retention was 2.20 times higher among mobile phone group (OR 2.20, 95%CI: 1.55 – 3.13, I^2^ = 53.20%, P < 0.001) compared to HIV-infected pregnant women in the control group (Fig 4). Pooled odds of ART uptake in HIV-infected pregnant women assigned with mobile phone interventions was 1.5 times higher than in the control group (OR 1.5, 95%CI: 1.07-2.11, I^2^ =0%, P = 0.020) (Fig 5). Evidence has shown statistically significant impact of mobile phone communication on HIV testing at 6 weeks and above among HIV exposed children (OR 1.89, 95%CI: 1.04 – 3. 48, I^2^ =88.04%, P = 0.032) (Fig 6). In the outcome Infant HIV infection among HIV-infected pregnant women, we found no statistically significant difference between the mobile phone communication and control groups (OR 0.81, 95%CI: 0.14 – 4.88, I^2^ = 62.18%, P = 0.821) (Fig 7). The meta-analysis of maternal ART adherence greater than 95% included only one study. The difference between the two groups was not statistically significant (OR 1.03, 95%CI: 0.07 – 1.50, P = 0.864) (Fig 8). Lastly, the pooled results of maternal viral load non-suppression included only one study. The difference between the mobile phone and control groups was not statistically significant (OR 0.88, 95%CI: 0.48 – 1.62, P = 0.692) (Fig 9).

**Figure 4:**
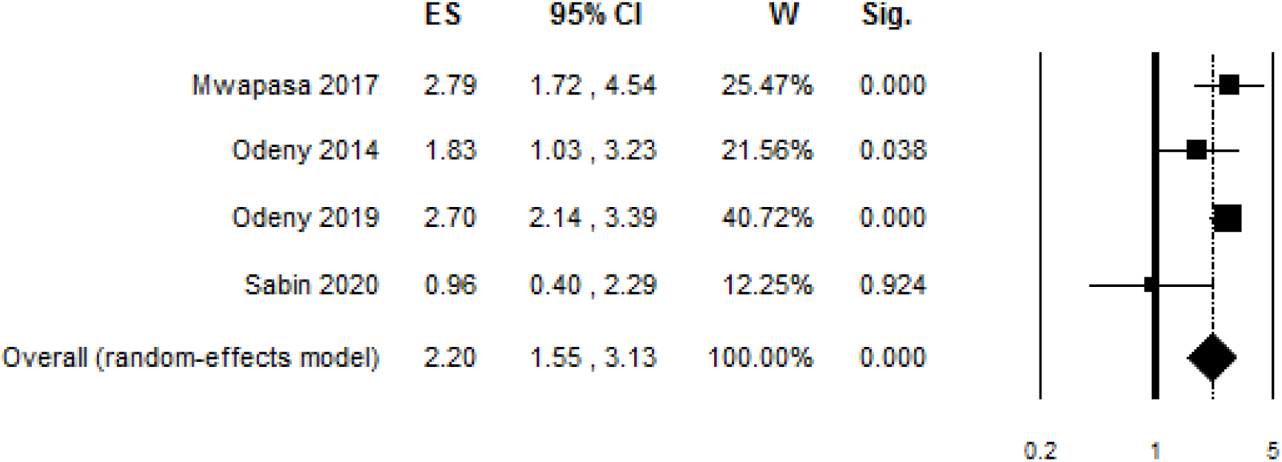
Forest plot of post-partum retention among HIV-infected pregnant women: mobile vs control groups

**Figure 5:**
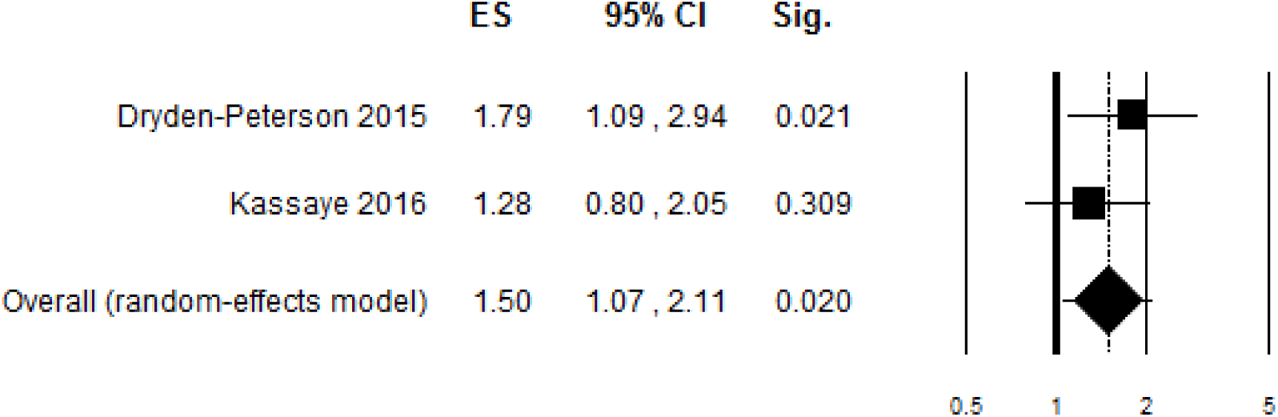
Forest plot of ART uptake among HIV-infected pregnant women: mobile phone vs control groups.

**Figure 6:**
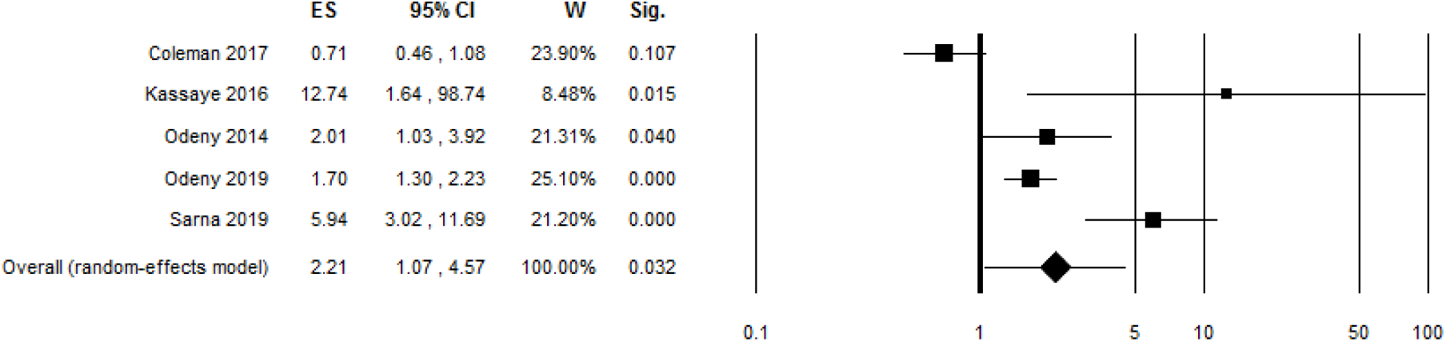
HIV testing at 6 weeks and above among HIV exposed children: mobile phone vs control groups.

**Figure 7:**
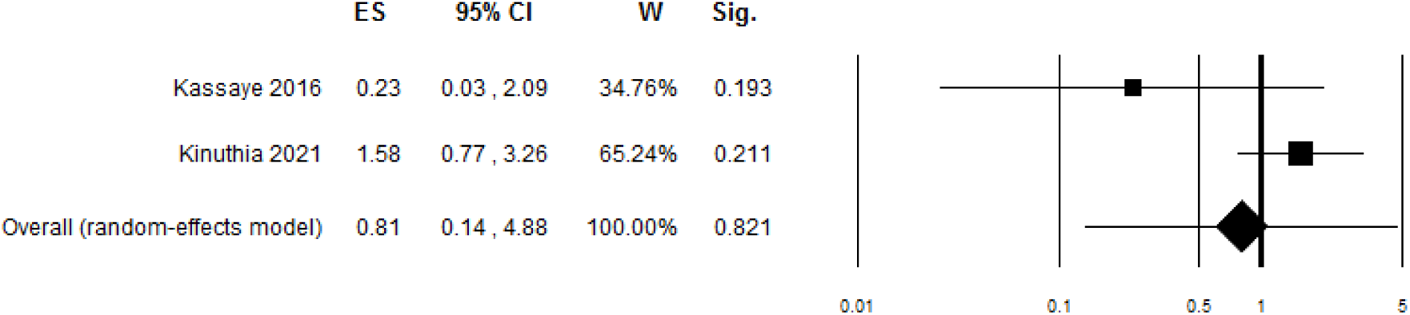
Infant HIV infection among HIV-infected pregnant women: mobile phone vs control groups.

**Figure 8:**
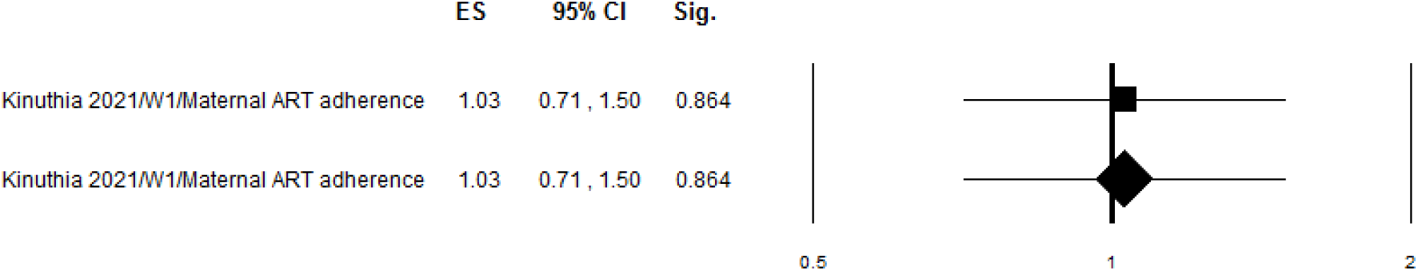
Maternal ART adherence >=95%: mobile phone vs control groups.

**Figure 9:**
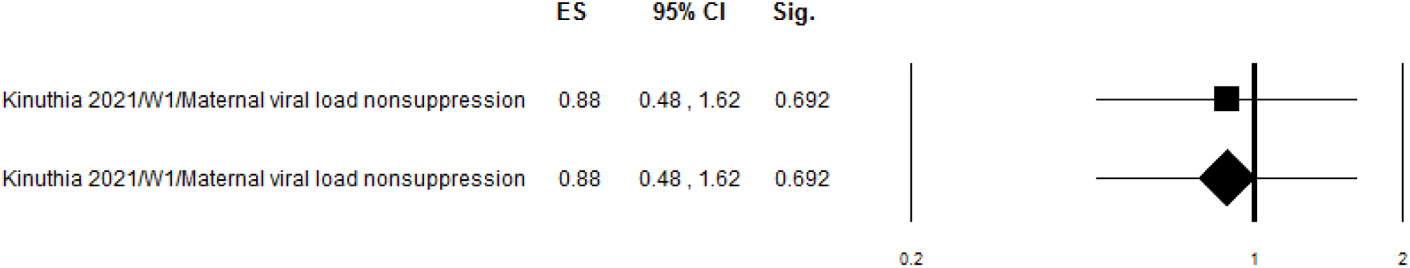
Maternal viral load non-suppression: mobile phone vs control groups.

**Figure 10:**
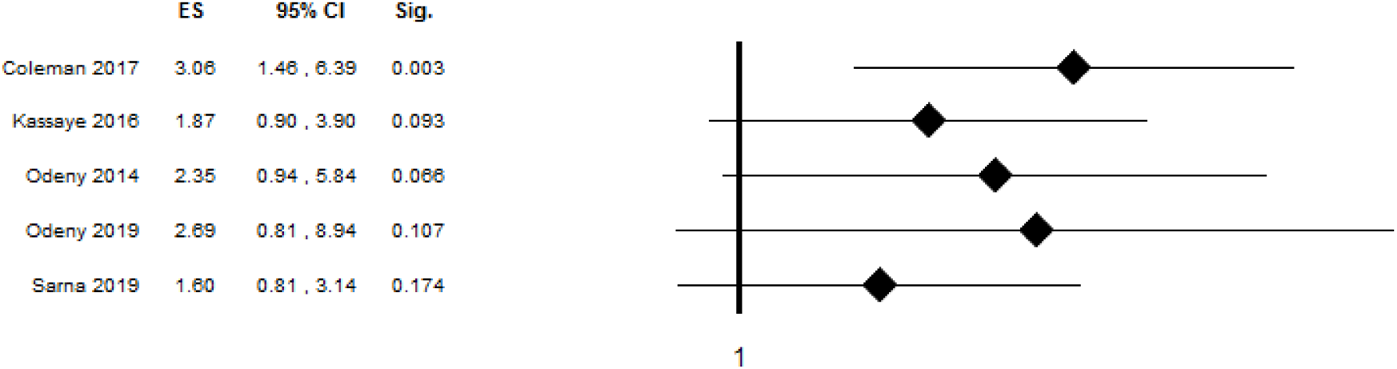
Sensitivity analysis of HIV testing at 6 weeks in children

### 3.4. Sensitivity analysis

To assess the robustness of our pooled OR of HIV testing at 6 weeks in children, we performed a sensitivity analysis. Sensitivity analysis revealed that the overall estimate of HIV testing in children at 6 weeks was not robust and was influenced by a single study [50]. The ORs for overall HIV testing in children at 6 weeks were 3.06 (95%CI: 1.46–6.39) [50] and 1.60 (95%CI: 0.81–3.14) [48], respectively (Fig 9).

## 4. Discussion

We conducted a systematic review and meta-analysis to determine the impact of mobile phone on 6, 830 HIV-infected pregnant women and 4, 111 HIV-exposed children. Our review found that mobile phone had a minor impact on ANC visits among HIV-infected pregnant women (RR = 1.02, 95 percent CI: 0.95 – 1.10, 1, 778 participants, 5 studies) [45, 48–50, 52]. In comparison to our previous review, which found that mobile phone communication was ineffective in improving ANC visits (RR 1.66, 95%CI 1.02 to 2.70, 381 participants, 1 study) [33], this current review confirmed the previous findings on ANC visits. The pooled post-partum retention study, on the other hand, included four studies [46–48, 52]. Post-partum retention was found to be 2.20 times higher in the mobile phone group than in the control group (OR 2.20, 95%CI: 1.55 – 3.13, P 0.001). Our post-partum retention findings are highly statistically significant and may be preferable to other post-partum retention interventions. The trial discovered that integration of care can lead to better transitions between prevention of mother-to-child transmission (PMTCT) and ART, as measured by a higher percentage of ART-eligible women starting ART within 12 months of testing HIV-positive in ANC in integrated care clinics versus non-integrated care clinics (40% vs. 17%; OR 3.22; 95%CI: 1.81–5.72) [53]. Previous reviews have found that text messaging is effective in increasing adherence to ART [53–56]. Finitsis et al. [57] reported a pooled OR of 1.48 (1.09 to 2.01) on any HIV outcome, but objective and subjective outcomes were pooled across all types of intervention as long as they included some text messaging. Similarly, in a randomized controlled trial that examined the use of monetary incentives to increase postpartum ART retention, the effect of conditional cash transfers on retention in PMTCT at six weeks postpartum was 12.5 percent higher probability of retention at six weeks postpartum for women in the intervention arm compared to the control arm (81% vs. 72%; RR ratio: 1.11; 95%CI: 1.00–1.24) [56]. Another individual randomized control trial (IRT) discovered that task-shifting to nurses, as well as home visits by peer counsellors to defaulted patients, did not result in a change in post-partum retention in the first 12 months after ART initiation [58].

In terms of the pooled odds of ART uptake in HIV-infected pregnant women, we found that mobile phone increase ART uptake when compared to our previous review, which found no statistically significant difference [33]. Similarly, our evidence has shown a statistically significant impact of mobile phones on HIV testing at 6 weeks and above among HIV-exposed children, which was also confirmed in our previous review [33]. In comparison to the first review, the updated version included a quite large number of included studies. Furthermore, maternal ART adherence, viral load non-suppression, and infant HIV infection were not statistically significant between intervention and control groups, which could be attributed to the small number of studies included in those outcomes. Our review has several clinical implications. Our findings, in particular, highlight the importance of mobile phone communication as an effective intervention for improving outcomes in HIV-infected pregnant women and their children during the postnatal period. However, more research is needed in the prenatal period. Another advantage of this review is that HIV-infected pregnant women are known to be a vulnerable study population, with poor ART outcomes and being understudied. In comparison to the previous review [33], this review included a much larger number of HIV-infected pregnant women. Because of its low cost, mobile phone intervention is a “low-hanging fruit” that PMTCT programs around the world could easily adopt and spread more broadly [47].

These results should be considered in a context of several limitations in included studies. All studies were conducted in sub-Saharan Africa, and the findings are likely to be an application in other similar settings. Due to the wide variation in interventions, study designs, and outcome measures, other forests included only one study in the meta-analysis [59]. The risk of bias is another limitation. Only two studies out of nine had a low risk of allocation concealment [45, 46] and also, two studies had low risk of participant and personnel blinding [45, 51]. Inadequate allocation concealment and participants and personnel blinding are influential RCT methodological factors. This may impact on the review validity. By the way, the overall strength of the evidence has been rated as moderate. Other important limitation of our study include may include the lack of electricity for charging phones in rural areas. A study cited a lack of electricity as a potential barrier to an mHealth intervention [59, 60]. Mobile phone-based interventions pose an added burden on daily battery consumption and policy makers should be aware of this challenge, particularly in rural areas and informal settlements, where electricity scarcity may hinder equitable access to the benefits offered [59]. Our findings may not be generalizable to women who never attended the clinic or do not use phones, and that more research is needed to quantify smartphone penetration in this and other low-resource areas. While phone ownership and usage preferences may differ in other contexts, phone use is rapidly increasing, and we sought to investigate user behaviour in this context to inform future interventions. Despite the fact that cell phone coverage has increased dramatically across the world, 477 million people in Sub-Saharan Africa subscribed to mobile services, accounting for 45% of the population at the end of 2019 [61]. Thus, many women do not have access to cell phones and thus could not benefit mobile phone interventions. Lastly, a more direct measure of adherence, such as therapeutic drug monitoring or antenatal and postnatal viral load monitoring, should be included in future studies.

## 5. Conclusions

Our findings can help to inform evidence-based decisions about how to improve outcomes for HIV-infected pregnant women and their children. This study found that using a mobile phone improved post-partum retention, ART uptake, and HIV testing at 6 weeks and above in HIV-infected pregnant women. However, no statistically significant results were found for ANC visits, maternal ART adherence, viral load non-suppression, or infant HIV infection. Given the growing importance of elimination of mother-to-child transmission (eMTCT) and the HIV-related impact in pregnant women, our research is critical.

More research is needed to determine the effect of mobile phones on prenatal period, maternal ART adherence, viral load non-suppression, and infant HIV infection. In light of the bias, our evidence has been rated as moderate. Further, the implementation of these findings should also consider the various barriers to phone ownership and coverage in various settings.

## Data Availability

All data produced in this present work are contained in the manuscript.

## Abbreviations

ANC: ante-natal consultation
ART: antiretroviral therapy
CENTRAL: Cochrane Central Register of Controlled Trials
CRTs: cluster randomized control trials
EACS: European AIDS Clinical Society
eMTCT: elimination of mother-to-child transmission
HAART: Highly active antiretroviral therapy
HIV: human immunodeficiency virus
IAS: International AIDS Society Conference on HIV Pathogenesis and Treatment
IRTs: individual randomized control trials
MeSH: Medical subject heading
mHealth: mobile health
MMWR: Morbidity and Mortality Weekly Report
MTCT: mother–to– child HIV transmission
ORs: Odds ratio
PICOS: Participants-Intervention-Comparisons-Outcomes-Settings
PCR: polymerase chain reaction
PMTCT: Prevention of mother-to-child transmission
PRISMA: Preferred Reporting Items for Systematic Reviews and Meta-analyses
RR: risk ratio
SMS: Short Message Service
UNAIDS: The Joint United Nations Programme on HIV/AIDS
US: United States
WHO: World Health Organization

## Acknowledgments

None

**Ethics approval and consent to participate:** Not applicable.

**Consent for publication:** Not applicable.

**Competing interests:** The authors have no competing interests to declare.

**Authors’ contributions:** The study was conceived and designed by JLT and CIO. JLT registered the review in Prospero and searched for studies. The data extraction was carried out by both JLT and PB. JLT carried out the study analysis and wrote the manuscript. All of the writers helped to edit the manuscript and approved the final version.

**Additional information:** All the review material has been included in this manuscript.

